# Ticagrelor Compared to Clopidogrel in aCute Coronary syndromes (TC4) – A Bayesian pragmatic cluster randomized controlled trial

**DOI:** 10.1101/2024.11.06.24316875

**Authors:** Stephen A. Kutcher, Nandini Dendukuri, Sonny Dandona, Lyne Nadeau, James M. Brophy

## Abstract

**Background:** Dual-antiplatelet therapy (DAPT) is the standard of care for acute coronary syndromes, but uncertainty exists regarding the optimal regime for North American patients.

**Methods:** This pragmatic, open-label, time clustered, randomized trial (ClinicalTrials.Gov (NCT04057300) compared the effectiveness and safety of DAPT with ticagrelor or clopidogrel in acute coronary syndrome patients from a single tertiary academic center in Montreal, Canada. The primary effectiveness endpoint was a composite of all-cause mortality, non-fatal myocardial infarction, or ischemic stroke. The primary safety endpoint were bleeding hospitalizations. Twelve-month outcomes were ascertained from the Québec universal electronic health databases. The study was designed and analyzed within a Bayesian paradigm to supplement existing knowledge. The primary analysis was a Bayesian logistic regression models with an informed focused prior from previously randomized North American patients. Robustness was evaluated with vague and other pre-specified informative priors, spanning reasonable pre-existing beliefs. Clinically significant benefits and harms were defined as risk reductions exceeding a 10% difference.

**Results:** 1,005 ACS patients were randomized to ticagrelor (n = 450) or clopidogrel (n = 555). MACE occurred in 50 (11.1%) ticagrelor and 64 (11.5%) clopidogrel patients (relative risk (RR), 0.95; 95% credible interval [95% CrI]: 0.67, 1.35 with a vague prior). The primary analysis with an informed focused prior resulted in probabilities of a clinically meaningful ticagrelor benefit (RR<0.9), equivalence (0.9 ≦ RR ≧, 1.1) or harm (RR ≧, 1.1) of 2%, 41% and 57%, respectively. For the safety endpoint, there was no consistent signal of benefit or harm with ticagrelor. Sensitivity analyses with a range of prior beliefs gave generally consistent results.

**Conclusions:** Whether this trial was analysed with a vague, or a range of reasonable informed priors, no strong evidence for the superiority of ticagrelor over clopidogrel was found.

## INTRODUCTION

Dual-antiplatelet therapy, the combination of a P2Y_12_ receptor inhibitor and acetylsalicylic acid (ASA), is the contemporary treatment strategy for the prevention of secondary ischemic-related events in patients with an acute coronary syndrome (ACS). According to clinical guidelines^1–3^ the P2Y_12_ inhibitor, ticagrelor, is favoured over clopidogrel. The strength of this endorsement is founded on the comparative Platelet Inhibition and Patient Outcomes (PLATO) trial^4^ performed in 862 study centers across 43 countries that reported a 16% reduction (hazard ratio (RR) 0.84, 95% confidence interval [CI] 0.77 - 0.92; P<0.001) in major acute cardiovascular events (MACE) with ticagrelor without any excess in major bleeding events.

However, PLATO effectiveness varied across the study regions as the 1,814 North American (NA) patients showed a statistically non-significant increase in MACE (RR_NA_ 1.25, 95% CI 0.93 - 1.67)^4,5^ with ticagrelor and a statistically significant deviation from the overall PLATO pooled result (p = 0.045). A Bayesian hierarchical (random effects) model accounting for the regional variability, provides a compromise between the pooled and individual geographical rates by a structured borrowing of information. Under this model, the PLATO NA ticagrelor risk estimate moves toward the PLATO global mean (RR_NA_ =1.13 (95% CI: 0.75 - 1.47)) but nevertheless leaves clinically important uncertainty about ticagrelor benefit in NA patients.^6^ Readers may have different prior beliefs about which data to consider or which statistical model best describes the underlying data generating mechanism, but if these different beliefs produce different inferences then the data cannot be considered robust. This provided the justification for additional randomized clinical trial (RCT) data to resolve the uncertainties about the comparative effectiveness of DAPT in a NA ACS population.

## METHODS

### Trial design and randomization

The **T**icagrelor **C**ompared to **C**lopidogrel in a**C**ute coronary syndromes – the TC4 trial (<u>NCT04057300</u>) – was a pragmatic, open-label, active control, time clustered RCT designed to assess the effectiveness and safety of ticagrelor versus clopidogrel as DAPT therapy in a ACS population undergoing percutaneous coronary intervention (PCI) in a single tertiary academic center in Montreal, Canada, between October 2018 and March 2021. Randomization followed a cluster randomized time-crossover design.^7^ In summary, all ACS patients arriving to the McGill University Health Center during a 2-month cluster would receive the scheduled DAPT for that period. Drug assignment for the first exposure period was determined by a random number with simple alternation for subsequent 2-month drug assignments periods. The COVID-19 pandemic severely hampered recruitment efforts in 2020, particularly disrupting ticagrelor recruitment clusters and consequently we added an extra 2-month ticagrelor recruitment period before study end. Given the large number of potential healthcare physicians (emergency room, cardiac catheterization, cardiology consultation, and coronary care unit) involved with ACS care, this time clustering randomization simplified administrative hurdles and rendered the trial feasible. Given the acute population studied, the unblinded exposure assignment was not expected to bias results.

### Trial oversite

The trial was investigator initiated, sponsored by a public funder (Canadian Institute of Health Research (reference number PJT-156344)), and approved by the research ethics board at the participating hospital. The 2018 funded and REB trial protocol is available online (https://www.brophyj.com/upload/TC4protocol.pdf). The trial follows the CONSORT reporting checklist (see Supplemental file). All statistical code is freely available upon request but ethical restrictions from the Quebec provincial health authorities prohibit the sharing of individual patient level data.

### Patients

Consecutive ACS patients admitted to the MUHC emergency department, cardiology or intensive care services between October 1, 2018, and March 31, 2021, and undergoing cardiac angiography were approached to participate. Operationally, this was achieved by continuous daily examination of the cardiac catheterization roster. Specifically, ST and non-ST elevation myocardial infarction (STEMI and NSTEMI) patients with positive biomarkers and patients with a diagnosis of unstable angina were eligible for study inclusion. A research nurse obtained informed consent following confirmation from the treating physician that DAPT was the appropriate treatment. Exclusion criteria were minimal and included i) lack of patient specific clinical equipoise by the attending physician ii) patient refusal iii) prior intolerance to either drug iv) recent (< 30 days) previous ACS hospitalisation v) out of province residency.

### Exposure

Patients were randomized to receive ticagrelor or clopidogrel depending on the date of their ACS hospitalization. In accordance with guidelines^1–3^ and previous trials^4^, physicians were advised to prescribe a 180 mg loading dose followed by a 90 mg bid dose of ticagrelor or a 300 mg loading dose followed by a daily dose of 75 mg of clopidogrel to their patients. Both therapies were to be accompanied by a 325 mg loading dose and 81 mg daily dose of aspirin, with patients encouraged to take their medications for 12 months following hospital discharge. Beyond the choice of DAPT regime, all other treatment decisions were made by treating physicians, independently of this study.

### Outcome

Similar to the PLATO trial^4^, the primary effectiveness measure was a 3-point MACE outcome – a composite of all-cause mortality, non-fatal MI (ICD-10 codes I21.X, I22.X, I23.X, I25.2), or ischemic stroke (ICD-10 codes H34.1, I63.X, I64.X, I67.X) – within 12 months of the index ACS hospitalization. The primary safety outcome was a composite of gastrointestinal bleeds (ICD-10 code K92.X) or hemorrhagic strokes (ICD-10 codes I60.X, I61.X, I62.X) requiring hospitalization. The secondary outcomes of interest were the individual MACE and safety outcomes. Patient follow up was monitored electronically via Québec medico-administrative hospital databases using ICD-10 codes and death certificates from hospital and provincial health records, which have been previously validated.^8–10^ Outcome data extraction was performed blinded to treatment assignment.

### Bayesian statistical design

In addition to specifying type I and type II error rates, frequentist sample size methods depend on point estimates of the intervention and control inputs including standard deviations, which are typically not accurately known at the study design stage. Bayesian methods offer several advantages including i) input uncertainties being represented by prior densities rather than point estimates with resolving uncertainty through the progressive accumulation of new evidence ii) this incorporation of existing prior knowledge with new data follows the laws of probability (via Bayes’ Theorem) iii) the resulting posterior distributions allow multiple direct probability statements to be formulated about clinically meaningful benefits, harm or practical equivalency iv) statistical penalties are not required for Bayesian sequential data analyses as posterior probabilities computed at the moment of stopping the trial are perfectly calibrated^11^.

Given the existence of previous, but inconclusive, evidence regarding the choice of DAPT in NA ACS patients^4,5^ this Bayesian study was designed to leverage that best available prior evidence with new data from the current study allowing the refinement of both efficacy and safety estimates with a reduced sample size compared to standard non-Bayesian designs. Using the PLATO^4,5^ regional NA cohort as an informed focused prior (RR_NA_ 1.25, 95% CI 0.93 - 1.67) and assuming the same PLATO NA event rates, we calculated a 1,000 patient study would result in a 96% posterior probability of an increased ticagrelor risk and an 83% probability that the difference exceeds an absolute 1% MACE difference. Since we believed these projected changes would be clinically important, our study was designed to reach this sample size. In summary, we believed adding 1,000 randomized subjects to the existing 1,800 randomized NA patients from PLATO was a justifiable and clinically meaningful addition to the evidence base.

### Bayesian priors

Given that our projected sample size was limited by available funding and was calculated by incorporating existing prior evidence, it is natural and appropriate that our analyses were also prespecified Bayesian methods updating prior beliefs with the current data. The resulting posterior distribution is a weighted mean of the prior and new data. The probabilities for clinically meaningful harm, benefit or equivalence are directly proportional to the area under the posterior probability density.

Bayesian analyses with both vague (enabling current study data to dominate) and informative priors are extremely useful in most studies but especially those with modest sample sizes. Priors have historically been considered the Achilles heel of Bayesian analyses, but this may be mitigated by formally acknowledging that clinical beliefs do vary,^12^ and consequently considering a community of priors which provides a test of robustness. It is worth recalling that frequentist models also involve numerous assumptions, often considerably more latent than those of explicit Bayesian priors.

The priors considered were a “vague” prior represented by a Student-t distribution around the null effect (RR=1.0), with 3 degrees of freedom and a sd of 5 and three informative priors. The three informative priors considered were i) a “focused” prior* (defined relative to our study population) from the NA PLATO MACE and bleeding estimates (RR_MACE_ = 1.25, 95%CI 0.93 - 1.67; RR_Bleed_ = 1.05, 95%CI 0.76 - 1.45) ii) an “enthusiastic” prior^†^ (defined relative to previous ticagrelor benefit) using the overall PLATO MACE (RR_MACE_ = 0.84, 95% CI 0.77 - 0.92) and bleeding estimates (PLATO Bleeding RR_Bleed_ = 1.04 95%CI 0.95–1.13), and iii) a “summary” prior^12^ elicited from a Bayesian Network Meta-analysis of 17 previously published RCTs involving 57,814 subjects ^13^ (RR_MACE_ = 0.95, 95%CrI: 0.81 - 1.14; RR_Bleed_ = 1.07, 95%CrI 0.99 - 1.17). In addition to these population level treatment effect priors, non-informative priors were used for the nuisance parameters, a Student-t prior, with 3 degrees of freedom (student_t(3, 0, 2.5) for the standard deviation (sd) and a Lewandowski-Kurowicka-Joe uniform distribution (η=1) for the correlation structure of the cluster-levels.

### Statistical analyses

The baseline characteristics of study participants were summarized using means, standard deviations, for continuous variables and proportions for categorical groups. Analyses examined the time to the first occurrence of a previously specified outcome with censoring at 1-year post randomization.

Given the use of the universal health care records there were no loses to follow-up. As no differences were noted between survival and logistic analyses, we report herein results from the logistic models. We also fit both fixed effect (pooled sample) and hierarchical models^14^ that account for the time clustering randomization.

Bayesian analyses provide posterior distributions for each parameter thereby providing estimates not only of their mean but also of their variability, commonly expressed as credible (CrI), or probability intervals. Consequently we are able to evaluate not only the probability of one treatment being better than the other but also the probability exceeding the smallest change in a treatment outcome that a patient, a care provider, or both would consider worthwhile, often referred to as the minimal clinically important difference (MCID).^15^ While these thresholds are admittedly arbitrary, they can be used to enhance the appreciation of the posterior treatment distributions. For example, we report MCIM probabilities statements based on a ±10% change in the relative risks as well as the probability for the range of practical equivalence (ROPE), where the posterior relative risks fall between 0.9 and 1.11. The full posterior distributions naturally permit the evaluation of differing clinical thresholds Bayesian statistical inference was performed using the brms R package^16^, a front end wrapper for the Stan probabilistic programming language^17^ which samples posterior distributions with the No-U-Turn Sampler (NUTS)^18^, an extension of the Hamiltonian Monte Carlo (HMC) algorithm. Three HMC chains, each a minimum of 10000 iterations with a 5000 burn-in period were used to produce a total of 15000 posterior samples. All samples were monitored to ensure their convergence. Model comparisons were done using the leave-one-out cross-validation (LOO), to evaluate model fit.^18^ The LOO cross-validation evaluation suggested the pooled model, when using a vague prior, had a better overall fit to the data than the hierarchical model. Consequently, unless otherwise specified results refer to this pooled model.

All analyses were performed within the RStudio integrated developmental environment^19^ using the R programming language^20^ and have followed Bayesian Analysis Reporting Guidelines^21,22^

## RESULTS

Between October 1, 2018, and March 31, 2021, 1,005 ACS patients were randomized and analyzed (Figure 1) from thirteen 2-month (7 clopidogrel and 6 ticagrelor) and one 4-month ticagrelor cluster periods (see Methods - Trial design and randomization). Follow-up, using the provincial electronic healthcare databases, ended in April 2022. The clopidogrel (N=555) and ticagrelor (N=450) DAPT groups were generally well balanced across their baseline characteristics (Table 1) with an average age of 67 and 75% males. All subjects had a 12-month follow-up unless a terminating outcome occurred prior to this time (Figure 2).

**Figure 1:**
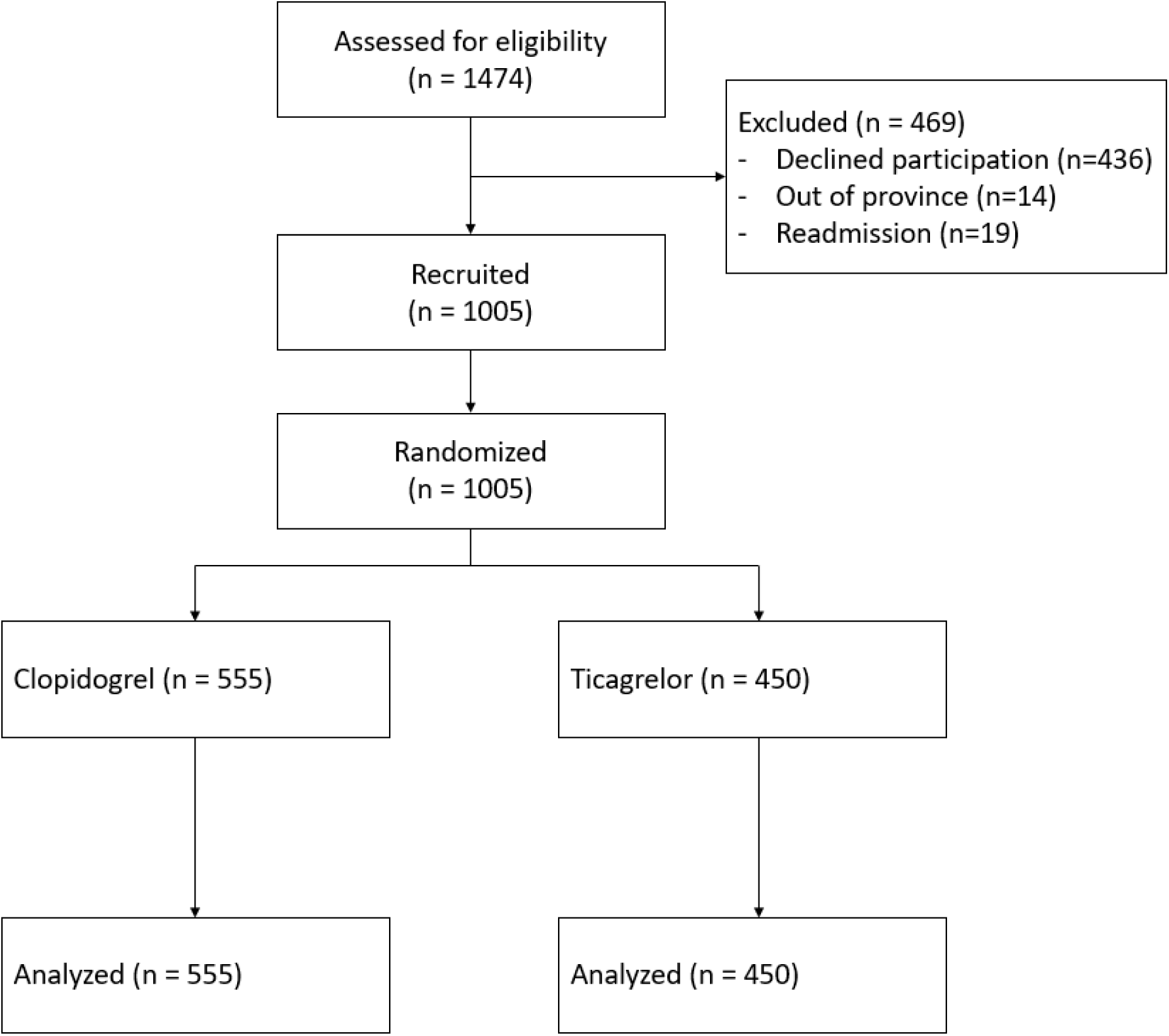
Flow chart of the TC4 study subjects.

**Figure 2:**
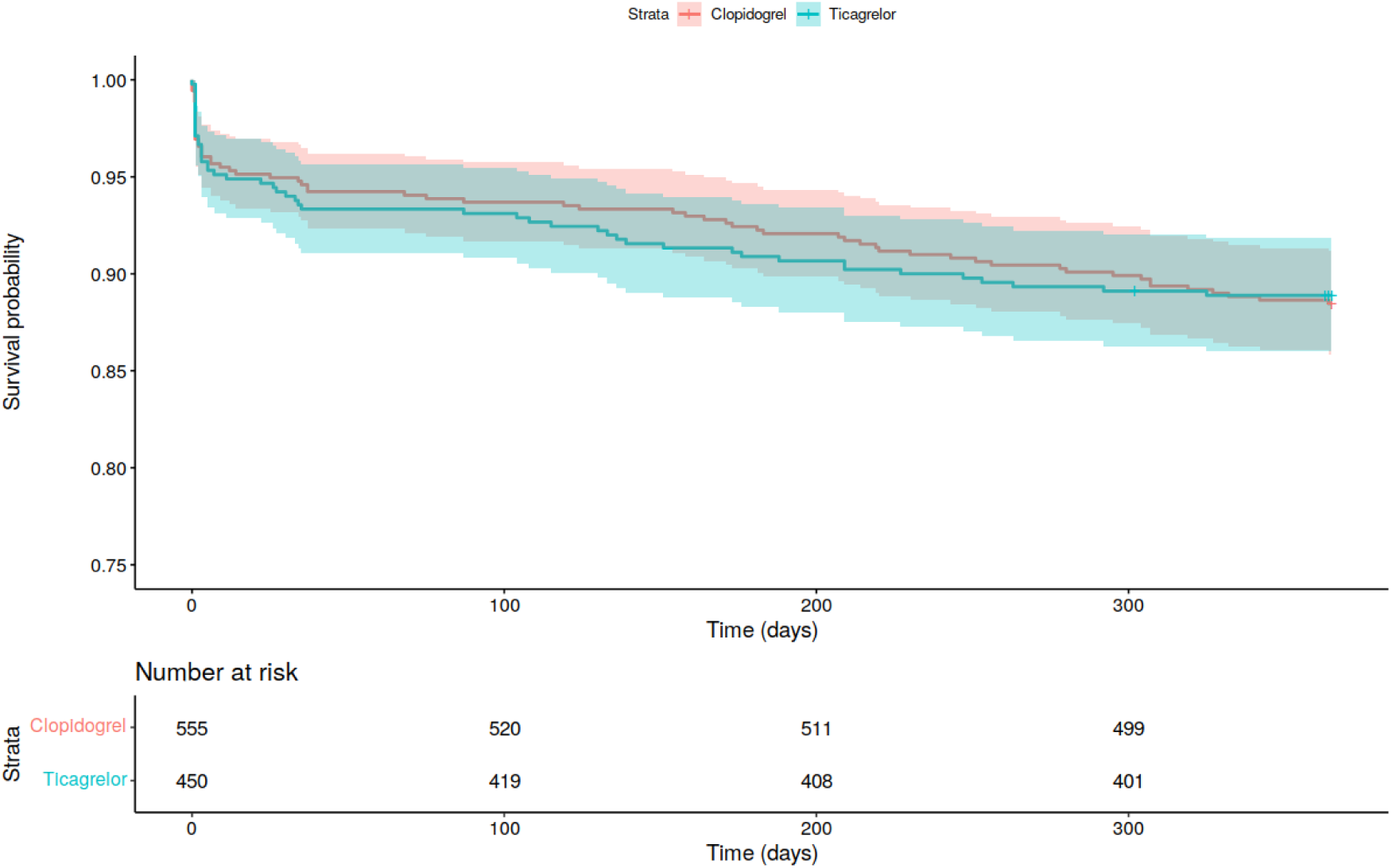
Kaplan Meier MACE curve for TC4 data.

**Table 1:** Baseline characteristics of the TC4 study population by DAPT treatment.

|  | Clopidogrel | Ticagrelor |
| --- | --- | --- |
| n | 555 | 450 |
| Age (mean (SD)) | 67.5 (10.9) | 65.2 (11.32) |
| Sex (male), n (%) | 420 (75.7) | 338 (75.1) |
| Height, cm (mean (SD)) | 170.6 (9.5) | 171.0 (9.3) |
| Weight, kg (mean (SD)) | 83.1 (22.0) | 83.3 (17.8) |
| Smoking status, n (%) |  |  |
| Never | 192 (35) | 181 (40) |
| Former, daily | 224 (41) | 157 (35) |
| Current, daily | 134 (24) | 110 (25) |
| Race, n (%) |  |  |
| Caucasian | 453 (81.6) | 376 (83.6) |
| Other | 102 (18.4) | 74 (16.4) |
| Previous DAPT, n (%) |  |  |
| None | 409 (74.1) | 341 (76.3) |
| Clopidogrel | 137 (24.8) | 88 (19.7) |
| Ticagrelor | 6 (1.1) | 17 (3.8) |
| Prasugrel | 0 (0.0) | 1 (0.2) |
| ACS diagnosis, n (%) |  |  |
| STEMI | 116 (20.9) | 94 (20.9) |
| NSTEMI | 210 (37.9) | 207 (46.1) |
| Unstable Angina | 228 (31.2) | 148 (33.0) |
| Hypertension, n (%) | 387 (69.9) | 300 (67.0) |
| SBP (mean (SD)) | 140.6 (22.2) | 140.0 (22.6) |
| DBP (mean (SD)) | 79.7 (13.7) | 80.4 (15.0) |
| Heart rate (mean (SD)) | 72.9 (15.4) | 72.4 (15.1) |
| Dyslipidemia, n (%) | 376 (68.0) | 301 (67.2) |
| Diabetic, n (%) | 185 (33.5) | 139 (31.0) |
| Type II, n (%) | 168 (90.8) | 130 (93.5) |
| Previous MI, n (%) | 159 (28.6) | 120 (26.9) |
| Previous PCI, n (%) | 144 (25.9) | 114 (25.4) |
| CHF, n (%) | 32 (5.8) | 15 (3.3) |
| Previous CABG, n (%) | 77 (13.9) | 32 (7.1) |
| Previous stroke, n (%) | 27 (4.9) | 14 (3.1) |
| History of PAD, n (%) | 5 (0.9) | 2 (0.4) |
| Creatinine (median [IQR]) | 83.0 [71.0, 97.0] | 83.0 [71.0, 97.00] |
| COPD, n (%) | 97 (17.5) | 64 (14.3) |
| Troponin (median [IQR]) | 205 [16, 2440] | 416 [25, 2521] |
| Missing n (%) | 144 (26.5) | 85 (19.1) |

*Abbreviations:*
*SBP = systolic blood pressure,*
*DBP = diastolic blood pressure*
*CHF = congestive heart failure*
*(N)STEMI = (Non)ST elevation myocardial infarction*
*COPD = Chronic obstructive lung disease*

### Effectiveness outcomes

The TC4 MACE effectiveness outcome occurred at similar rates in both exposure groups (clopidogrel, 11.5%: ticagrelor, 11.1%) and the relative risk with a vague prior is 0.95 (95% CrI 0.67, 1.35) (Table 2). This equates to a 37% probability that ticagrelor is responsible for a clinically meaningful reduction (RR<0.9), a 21% chance of a clinically meaningful increase (RR>1.11), and a 42% probability being within the ROPE, when compared with clopidogrel (Table 2). While a vague prior means the posterior probabilities are completely dominated by the current TC4 data, it is evident from the original design and the spread of this posterior distribution that this analysis doesn’t allow for definitive conclusions.

**Table 2:** Results.

| Outcome | TC4 |  | Prior source (distribution) | Posterior probabilities |  |  |  |
| --- | --- | --- | --- | --- | --- | --- | --- |
|  | Clopidogrel<br>N=555 | Ticagrelor<br>N=450 |  | RR (95% CrI) | MCID benefit<br>(Pr<sub>HR<0.9</sub>) | ROPE<br>(Pr<sub>HR[0.9, 1.11]</sub>) | MCID harm<br>(Pr<sub>HR>1.11</sub>) |
| **MACE** | 64 (11.5%) | 50 (11.1%) | Vague<br>(t(3, log(1), 5)) | 0.95 (0.67, 1.35) | 0.37 | 0.42 | 0.21 |
|  |  |  | Focused<sup>1</sup><br>(N(1.23, 0.11)) | 1.12 (0.91, 1.39) | 0.02 | 0.41 | 0.57 |
|  |  |  | Enthusiastic<sup>2</sup><br>(N(0.84, 0.04)) | 0.85 (0.78, 0.93) | 0.89 | 0.11 | 0.00 |
|  |  |  | Summary<sup>3</sup><br>(N(0.97, 0.13)) | 0.99 (0.69, 1.38) | 0.30 | 0.44 | 0.26 |
| **All-cause mortality** | 15 (2.7%) | 7 (1.6%) | Vague<br>(t(3, log(1), 5)) | 0.60 (0.24, 1.39) | 0.83 | 0.09 | 0.08 |
|  |  |  | Focused<br>(N(1.24, 0.32)) | 1.05 (0.69, 1.58) | 0.24 | 0.35 | 0.41 |
|  |  |  | Enthusiastic<br>(N(0.79, 0.15)) | 0.75 (0.54 – 1.06) | 0.78 | 0.11 | 0.11 |
|  |  |  | Summary<br>(N(0.86, 0.13)) | 0.67 (0.27 – 1.52) | 0.76 | 0.11 | 0.13 |
| **MI** | 46 (8.3%) | 38 (8.4%) | Vague<br>(t(3, log(1), 5)) | 1.02 (0.67, 1.54) | 0.29 | 0.37 | 0.35 |
|  |  |  | Focused<sup>1*</sup><br>(N(1.24, 0.16)) | 0.99 (0.61 - 1.60) | 0.35 | 0.32 | 0.33 |
|  |  |  | Enthusiastic<br>(N(0.83, 0.05)) | 0.90 (0.81 – 1.01) | 0.48 | 0.52 | 0.0 |
|  |  |  | Summary<br>(N(0.96, 0.11)) | 1.05 (0.70 - 1.54) | 0.23 | 0.36 | 0.41 |
| **Stroke** | 3 (0.5%) | 5 (1.1%) | Vague<br>(t(3, log(1), 5)) | 1.99 (0.52 - 7.88) | 0.12 | 0.07 | 0.81 |
|  |  |  | Focused<sup>1*</sup><br>(N(1.73, 1.87)) | 1.93 (0.75 - 5.07) | 0.06 | 0.07 | 0.87 |
|  |  |  | Enthusiastic<br>(N(1.17, 0.18)) | 1.20 (0.93 - 1.55) | 0.01 | 0.24 | 0.75 |
|  |  |  | Summary<br>(N(1.02, 0.16)) | 1.11 (0.94 - 1.32) | 0.01 | 0.43 | 0.56 |
| **Bleeding** | 28 (5.0%) | 20 (4.4%) | Vague<br>(t(3, log(1), 5)) | 0.89 (0.52, 1.52) | 0.51 | 0.26 | 0.23 |
|  |  |  | Focused<br>(N(1.05, 0.23)) | 1.03 (0.78 - 1.36) | 0.17 | 0.51 | 0.32 |
|  |  |  | Enthusiastic<br>(N(1.04, 0.05)) | 1.03 (0.95 - 1.12) | 0.00 | 0.93 | 0.07 |
|  |  |  | Summary<br>(N(1.16, 0.16)) | 1.07 (0.99, 1.16) | 0.00 | 0.74 | 0.26 |

<sup>1</sup> The “focused” prior considers only the PLATO North American subjects (both Canadian and American).
<sup>1\*</sup> The “focused” prior for MI and stroke are based on PLATO US data alone as separate Canadian data could not be found so the priors for these outcomes.
<sup>2</sup> The “enthusiastic” prior considers only the PLATO trial data, the largest and most positive results in favor of ticagrelor. This is also considered “enthusiastic” as the standard deviation assumes no heterogeneity in the PLATO subpopulations according to geographic region of randomization. However, there is important regional variation that was been ignored in the original publication. Recognizing this variability, a PLATO hierarchical prior that acknowledges this regional variability can be determined. Using this hierarchical statistical model, the “enthusiastic” PLATO alone prior is modified and with an increase in the standard deviation as described by a N(0.84, 0.14) distribution<sup>6</sup> resulting in a posterior distribution RR = 1.03 95% CI 0.73 - 1.44. This “modified enthusiastic” prior results in an important shift for MCID MACE probabilities for ticagrelor benefit, equivalence and harm of 22%, 35%, 43%, respectively. “Modified enthusiastic” priors are not available for the other outcomes as PLATO regional outcome data was not located.
<sup>3</sup> The “summary” prior considers all randomized trials from a network meta-analysis of ticagrelor versus clopidogrel
BNMA = Bayesian network meta-analysis, MACE = major adverse cardiac events, MCID = minimally clinically important difference, MI = myocardial infarction, N = normal distribution, NA = North American, RR = risk ratio, ROPE = region of practical equivalence, t = t distribution

However, our prespecified primary Bayesian analysis was with a “*focused”* informative prior, based on the previously randomized PLATO NA subjects. This resulted in a posterior mean MACE RR = 1.12 (95%CrI 0.91, 1.39) with the probability of clinically meaningful ticagrelor effects (± > 10% change in RR) were 2% for benefit, 57% probability for harm, and 41% for clinical equivalence (see Table 2). These results are also displayed graphically in Figure 3, demonstrating that the posterior distribution is a weighted mean of the prior and new data from this trial. The probabilities for clinically meaningful harm, benefit or equivalence are directly proportional to the area under the posterior probability density.

**Figure 3:**
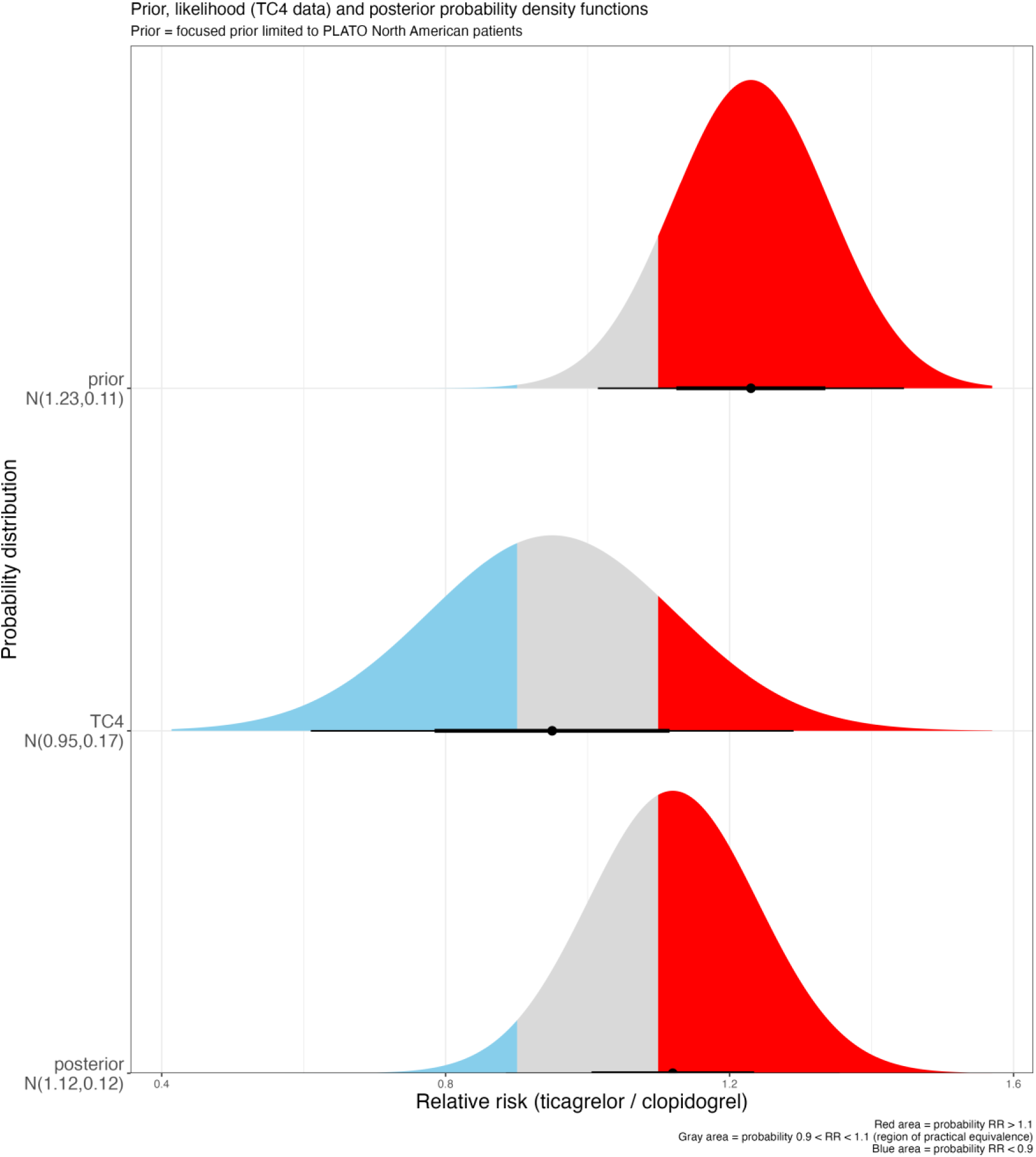

Posterior MACE probabilities employing the “*enthusiastic*” prior of all PLATO data shifted the posterior distribution to more favorable ticagrelor inferences with an 89% probability of a clinically meaningful improvement. However, as discussed below in detail, this “*enthusiastic*” prior is a very selective choice of the available prior RCT evidence. Posterior MACE probabilities with the “*summary*” prior, which was not restricted to the single PLATO study but rather included all high quality prior RCT evidence, gave less encouraging findings for any ticagrelor benefits (Table 2). The MACE posterior distribution mean with the “*summary”* prior was RR, 0.99 (95%CrI 0.69, 1.38) with a 30% probability of a clinically meaningful ticagrelor benefit but also a 26% probability of a clinically meaningful ticagrelor harm relative to clopidogrel and a 44% probability of clinical equivalency.

### Safety outcomes

With a vague prior, major bleeding events requiring hospitalizations did not substantially differ across the TC4 treatment groups (clopidogrel, 5.0%; ticagrelor, 4.4% (RR 0.89 (95%CrI 0.52 - 1.52). Expressed in term of clinically meaningful probabilities of ticagrelor to clopidogrel bleeding, showed a 51% probability for a reduction (RR<0.9), a 26% probability for clinical equivalence (0.9 < RR< 1.1), and a 23% probability for an increase (RR>1.11). As expected, conclusive safety inferences from this study alone is not possible. However, summarizing the integration of current trial bleeding events with the other priors (Table 2), showed only a very small chance of a clinically meaningful reduction in bleeding with ticagrelor (0-17%), a moderate to high probability of clinical equivalence (51-93%) and a modest probability of a clinically meaningful increase in bleeding rates (7-32%).

### Secondary outcomes

For the individual MACE components of TC4, there were fewer deaths from all-cause mortality identified in the ticagrelor group (1.6% vs. 2.7%) than the clopidogrel arm (RR 0.60, 95%CrI 0.24 - 1.39). No difference for MI events between ticagrelor and clopidogrel patients (8.4% vs. 8.3%, RR 1.02, 95%CrI 0.67 - 1.55). Lastly, a similar number of ischemic stroke outcomes between the two groups (clopidogrel, N=3 vs. ticagrelor, N=5) was observed (RR 1.99, 95%CrI 0.52 – 7.88). The power of this trial to make any meaningful inferences for these secondary outcomes is very limited. However, the primary analysis with the “*focused*” informative NA PLATO prior showed the probability of clinically meaningful harm with ticagrelor exceeded its probability of clinical benefit for all the individual MACE components.

## DISCUSSION

This pragmatic, time-clustered, registry trial successfully enrolled 1,005 ACS patients to ticagrelor or clopidogrel increasing by 60% the existing randomized evidence base of North American patients^4^ addressing the relative merits of the two different DAPT options following an acute coronary syndrome. Although our trial was not powered to detect meaningful outcome differences solely on its own merits, it was powered to assist in resolving existing uncertainties about DAPT choice in North American patients^4^. On its own (with a vague prior), this trial revealed approximately equal probabilities for a clinically important MACE benefit (37%), equivalence (42%), or harm (21%) with ticagrelor compared to clopidogrel. However, our primary prespecified analysis with an informative “*focused*” prior using PLATO North American data, markedly reduced the uncertainty regarding the relative efficacy of these two strategies in this population. Specifically, when this prior knowledge was updated with our current TC4 data, there was only a very small 2% probability of a clinically meaningful MACE benefit with ticagrelor compared to clopidogrel, and moderate probabilities of clinical equivalency (41%) or harm (57%) (see Table 2, Figure 3).

To account for other possible prior beliefs, our MACE results are also presented in combination with an “*enthusiastic”* prior, based on overall PLATO results^4^, and a systematic “*summary”* prior, based on a Bayesian network meta-analysis^13^. Using the “*enthusiastic*” prior, the probability of a clinically meaningful ticagrelor benefit was 89%. However, this prior minimizes uncertainty by ignoring the observed PLATO geographic regional variations. Using all the PLATO data, but accounting for the observed regional heterogeneity as a “*modified enthusiastic*” prior, the probability of a ticagrelor clinical benefit falls to 22% with a two-fold higher probability (43%) of clinical harm (see foot note of Table 2). Therefore, using a wide spectrum of reasonable priors, the probability of a clinically significant ticagrelor MACE benefit does not exceed 30% and may be as low as 2%. The MACE probability of clinical equivalence is approximately 40% and the probability of clinically significant harm between 20-50%. It is only with the sensitivity analysis using the highly selective “*enthusiastic*” prior of PLATO data alone (with a statistical model that underestimates uncertainty by neglecting between country variations) and ignoring the eight other prior comparative RCTs (n=8016)^13^ does the probability of clinically meaningful ticagrelor benefit approach an interesting level. Using the systematic “*summary”* prior, the probability of a clinically meaningful ticagrelor MACE benefit (RR < 0.9) is 30%, roughly equal to the probability (26%) of a meaningful increase in harm (RR >1.1). However, it should be noted that this “*summary*” prior suffers from the same over-estimation as the “*enthusiastic*” prior as PLATO data accounts for approximately 50% of the “*summary*” prior sample size.

Regarding the individual MACE components, our primary analysis with a “*focused*” prior showed little support for any ticagrelor superiority. There was a 24% probability of a clinically meaningful decrease, a 35% probability of practical equivalence and a 41% probability of a clinical meaningful increase in all-cause mortality. Similarly for the other secondary outcomes (MI or stroke) there were no clear signals of ticagrelor benefit with the “*focused*” prior, as the probability of clinically meaningful harm often exceeded that for benefit. As predicted at the planning stage, data analyses with a vague prior lacked the power to draw any meaningful inferences (Table 2). Analyses with the “*enthusiastic*” priors did suggest moderate probabilities for ticagrelor in reducing all-cause mortality and MI. However as noted above, these “*enthusiastic*” priors are at risk of bias and any ticagrelor benefits are attenuated with the less biased “*summary*” prior. Although the stroke numbers were small and inconclusive, all analyses independent of the chosen prior had posterior probabilities of a clinically meaningful increase in stroke exceeding 50%.

Unsurprisingly, the TC4 trial alone was underpowered to reliably identify clinically meaningful safety outcomes. With a vague prior, the probability of major bleeding events requiring hospitalization being reduced by a clinical meaningful benefit was 51% for ticagrelor compared with clopidogrel. There was also a 26% probability of equivalence and a 23% of a clinically meaningful bleeding increase with ticagrelor. When using informative priors, there was at least moderate probability (50-93%) of clinical equivalence compared to a smaller probability for increased ticagrelor harm (25%).

Ultimately, our TC4 trial findings, either when taken alone or integrated with a reasonable spectrum of prior beliefs do not fully align with North American ACS guidelines,^3,4^ which recommend ticagrelor over clopidogrel based on the single PLATO trial, a multinational study dominated by not by North American centres but by Western and Eastern European centres. TC4 trial efficacy results fall between the overall PLATO and its North American subgroup results and are compatible with the Bayesian network meta-analysis efficacy summary prior. Our results are also consistent with the recent comparative ALPHEUS trial^23^ of 1910 French and Czech patients randomly assigned ticagrelor or clopidogrel which found no difference their primary outcome, a composite of PCI-related outcomes at 48 hours (odds ratio [OR] 0·97, 95% CI 0·80–1·17; p=0·75)^‡^.

The heterogeneity between the overall PLATO results and the PLATO North American subgroup has been variously attributed to the play of chance^24^, a post hoc hypothesis whereby the higher NA aspirin doses exerted a negative interaction with ticagrelor^24^ or PLATO trial management irregularities^25–27^. The high dose aspirin hypothesis appears convoluted and there are several reasons why this may be seen as unlikely. First is the possibility of a Type 1 error, as this putative interaction was observed following multiple interaction testing of 46 expressions involving 37 factors^24^. Second in a large meta-analysis of antithrombotic trials^28^ comparisons of high versus low dose aspirin (n=10 RCTs, 6,767 subjects) or aspirin alone versus combined with other antiplatelet medications (n=27 RCTs, 34,452 subjects) there was no evidence for any differential dose impact on outcomes.

Regardless, given the observed PLATO heterogeneity, it is surprising that this is the first ticagrelor / clopidogrel ACS RCT, to our knowledge, to be performed in North America since the original 2009 PLATO publication^4^ and consequently represents a major study strength. We were able to recruit a large (N=1,005) sample of ACS patients achieving excellent baseline covariate balance across the DAPT treatment arms. The pragmatic nature of this trial, by limiting the exclusion criteria for patient participation, allowed for the investigation of the treatments in a more clinically representative population than a typical RCT. This may explain why the TC4 patients were older, on average than the PLATO study participants (∼67 years vs. ∼62 years). The TC4 study also recruited only those ACS subjects undergoing cardiac catheterization which is the typical North American ACS approach compared to only 64% in PLATO. By leveraging the provincial electronic healthcare databases, and focusing on clinically validated outcomes, we were also able to minimize the loss of patients during the 1-year follow-up period for identifying clinical outcomes. The Bayesian analytical approach eliminates null hypothesis significance testing and P-values.^5^ Instead, it allows for the incorporation of a wide-range of prior beliefs and permits formulation of direct probability statements regarding the effectiveness and safety evidence, and their interpretation with respect to clinically meaningful effect sizes. Finally, large comparative phase 3 or phase IV cardiovascular RCTs can cost approximately $33,000 per patient randomized.^29^ Our unique and innovative research design cost approximately $300 per patient randomized or less than 1% of typical RCT costs.

A limitation of the TC4 trial is its reliance on an intention to treat (ITT) analysis, ignoring any possible benefits from per protocol analyses.^7^ Unfortunately, the per-protocol and as-treated analyses were unachievable due to structural restrictions of the electronic follow-up databases whereby universal prescription follow-up data is only available for those subjects older than 65 years of age.

This pragmatic RCT added a substantial quantity of North American evidence to the DAPT ACS literature. As expected, the stand-alone TC4 results did not find conclusive evidence for the superiority of ticagrelor over clopidogrel for either the primary effectiveness or safety outcomes. After the incorporation of a range of clinically relevant priors, selected from the published literature, the results still do not demonstrate, in any convincing manner, the superiority of ticagrelor over clopidogrel for DAPT treatment. In fact, the probability of excessive ticagrelor harm is approximately equal to the probability of its benefit. Given these observations of a low probability for any ticagrelor efficacy superiority, its increased costs, and its inconvenience of twice daily dosing, it is difficult to accept guideline recommendations^1–3^ for choosing ticagrelor over clopidogrel, particularly for a North American patient population.

In conclusion, our study highlights the benefit of novel research designs and analyses to resolve ongoing uncertainties. Finally, given our findings and our conflicting interpretation of the evidence with the current clinical ACS guidelines, we suggest a comprehensive re-evaluation of DAPT choice for ACS patients should be considered.

## Data Availability

All statistical programs will be shared. Governmental regulations prohibit sharing patient level data.

https://www.brophyj.com/upload/TC4protocol.pdf

## Footnotes

* As noted in our 2018 funding protocol (https://www.brophyj.com/upload/TC4protocol.pdf), this prior was prespecified as our primary Bayesian analysis

† The “enthusiastic” prior considers only the PLATO trial data, the largest and most positive results in favor of ticagrelor. This summary prior ignores the PLATO geographic heterogeneity. Recognizing this variability, a PLATO hierarchical prior that acknowledges this regional variability can be determined^6^. Using this hierarchical statistical model, a “modified enthusiastic” PLATO prior with an increased standard deviation as described by a N(0.84, 0.14) distribution can and should also be considered.

‡ Due to the short duration of this trial it is not included in the Bayesian network meta-analysis which examines longer term outcomes.

